# Performance of progressive generations of GPT on an exam designed for certifying physicians as Certified Clinical Densitometrists

**DOI:** 10.1101/2023.07.25.23293171

**Authors:** Dustin Valdez, Arianna Bunnell, Sian Y. Lim, Peter Sadowski, John A. Shepherd

**Affiliations:** University of Hawaii Cancer Center; Hawaii Pacific Health Medical Group; University of Hawaii at Manoa; Univeristy of Hawaii Cancer Center

**Keywords:** GPT, LLM, clinical densitometry, CCD

## Abstract

**Background:** Artificial intelligence (AI) large language models (LLMs) such as ChatGPT have demonstrated the ability to pass standardized exams. These models are not trained for a specific task, but instead trained to predict sequences of text from large corpora of documents sourced from the internet. It has been shown that even models trained on this general task can pass exams in a variety of domain-specific fields, including the United States Medical Licensing Examination. We asked if LLMs would perform as well on a much narrower subdomain tests designed for medical specialists. Furthermore, we wanted to better understand how progressive generations of GPT (generative pre-trained transformer) models may be evolving in the completeness and sophistication of their responses even while generational training remains general. In this study, we evaluated the performance of two versions of GPT (GPT 3 and 4) on their ability to pass the certification exam given to physicians to work as osteoporosis specialists and become a certified clinical densitometrists.

**Methods:** A 100-question multiple-choice practice exam was obtained from a 3^rd^ party exam preparation website that mimics the accredited certification tests given by the ISCD (international society for clinical densitometry). The exam was administered to two versions of GPT, the free version (GPT Playground) and ChatGPT+, which are based on GPT-3 and GPT-4, respectively (OpenAI, San Francisco, CA). The systems were prompted with the exam questions verbatim. If the response was purely textual and did not specify which of the multiple-choice answers to select, the authors matched the text to the closest answer. Each exam was graded and an estimated ISCD score was provided from the exam website. In addition, each response was evaluated by a rheumatologist CCD and ranked for accuracy using a 5-level scale. The two GPT versions were compared in terms of response accuracy and length.

**Results:** The average response length was 11.6 ±19 words for GPT-3 and 50.0±43.6 words for GPT-4. GPT-3 answered 62 questions correctly resulting in a failing ISCD score of 289. However, GPT-4 answered 82 questions correctly with a passing score of 342. GPT-3 scored highest on the “Overview of Low Bone Mass and Osteoporosis” category (72% correct) while GPT-4 scored well above 80% accuracy on all categories except “Imaging Technology in Bone Health” (65% correct). Regarding subjective accuracy, GPT-3 answered 23 questions with nonsensical or totally wrong responses while GPT-4 had no responses in that category.

**Conclusion:** If this had been an actual certification exam, GPT-4 would now have a CCD suffix to its name even after being trained using general internet knowledge. Clearly, more goes into physician training than can be captured in this exam. However, GPT algorithms may prove to be valuable physician aids in the diagnoses and monitoring of osteoporosis and other diseases.

## Introduction

With the rapid evolution of machine learning and artificial intelligence (AI), many tasks in clinical medicine are now being automated with an accuracy that matches or exceeds human performance. Examples include the detection and classification of cancer from medical imaging [1-3], COVID-19 applications [4, 5], applications in Surgery [6], and the use of natural language processing (NLP) for interpreting medical records [7]. One form of NLP, large language models (LLMs) trained in an autoregressive manner on millions of documents, have gained a lot of attention due to their ability to take and, in many cases pass, certification examinations.

LLMs can be used to analyze, summarize, and generate human language that is meaningful and useful. Models in the GPT family are LLMs based on the Transformer artificial neural network architecture style and trained on millions of documents by OpenAI. ChatGPT is an instance of the GPT-3 model that has been further trained to be a helpful chatbot. The paid version, ChatGPT+, is a similar system that uses the larger GPT-4 model. Here we refer to the systems by their underlying architecture and release version, GPT-3 and GPT-4 [8, 9].

GPT-3 has shown an ability to perform well on different medical and professional certification examinations. As of February 12, 2023, GPT-3 has passed a multitude of examinations such as the Bar Exam[10], Stanford’s Medical School clinical reasoning final[11] and the United States Medical Licensing Examination [12, 13]. Through incorporating a wide variety of internet information into its training corpus, ChatGPT can answer clinically-relevant medical questions with fairly high accuracy, for example scoring 86.70% on a USMILE sample test [14]. Rarely is the passing of one exam the sole qualification for working within a field or subfield, but it is often used as an objective component in assessing one’s educational level. An example is the professional recognition of Certified Clinical Densitometrist (CCD) by the International Society for Clinical Densitometry (ISCD).

The ISCD CCD exam, introduced in 1996 [15], is a professional certification developed to recognize clinicians and professionals who have demonstrated entry-level knowledge and skill to interpret central bone density scans for the purpose of diagnosing osteoporosis. The exam is given around the world and is updated by ISCD on a regular basis. The exam is created from over 200 well-vetted Official Positions of the ISCD [16]. The exam is accredited by the National Commission for Certifying Agencies (NCCA) and consists of 125 multiple choice questions (100 graded questions + 25 experimental non-graded questions) and candidates have three hours to complete the examination. Besides have one of the following broad medical credentials; licensed MD (or DO, PAC, NP), Licensed Medical Practitioner Resident/Fellow or Non-licensed medical practitioner (PhD)), the CCD exam is the only other criterion necessary to gain the CCD certification. We know of no previous work using of an AI system to take the ISCD CCD exam.

We asked if GPT models, without augmented training in the specialty, could pass the CCD exam. Furthermore, we wanted to know if progressive versions of GPT score higher on the exam. In this study we evaluated the performance of two versions of GPT on their ability to pass the domain specific ISCD CCD test exam, and to evaluate the completeness and quality of their responses.

## Methods

The study is a simple cross-sectional survey design with two subjects. A full practice exam was obtained from a 3^rd^ party exam preparation website and fed to two versions of GPT, GPT-3 and GPT-4 (GPT playground and ChatGPT+, respectively). Each question of the exam was fed verbatim into both versions, and all responses were recorded. The responses were graded by the 3rd party exam website. The responses were also compared by category and reviewed by a rheumatologist with a CCD certification.

### Test Examination

The official CCD exam covers five major categories, with their approximate distribution as follows: Overview of Low Bone Mass and Osteoporosis (15%), Imaging Technology in Bone Health (23%), Clinical Application of Bone Densitometry (32%), Prevention and Risk Assessment (16%), and Treatment of Low Bone Mass and Osteoporosis (14%) [17]. A practice test was obtained from a 3^rd^ party examination website (examedge.com). The exam provider offers up to 10 online practice exams created from 1,000 unique questions and also allows you to select one of three exam modes (timed, untimed, study guide). We used the study guide option so each answer with explanation could be captured at the same time. The same test was administered to GPT-3 and GPT-4 using the reset option, which allows you to retake the test with the same questions. All questions were in multiple choice format and were copied verbatim into the GPT playground/ChatGPT+ prompt. An example question is, “The images acquired in a DXA scan of the lumbar spine will be used to check the patient’s: Bone mineral content. Nerve impulses. Tissue health. Blood pressure.” With the four multiple choice answers following the question in the order that the question listed them and separated with a period. All 100 questions were inputted, and their responses were recorded verbatim. There were two outputs generated by the test provider, a grade in terms of percent of questions right, and an ISCD score. The CCD exam has a possible score range of 150 to 400, as defined by the ISCD. To pass, you need a score of 300.

### GPT models

GPT-3 (version 3.0, released June 11, 2020) was accessed via GPT Playground[18] on March 20, 2023. GPT-4 (released March 14, 2023) was accessed through a paid subscription to OpenAI through the ChatGPT+ platform and accessed on March 20, 2023. A general overview of the differences between GPT-3 and GPT-4:

1. Model size: One of the most significant differences between GPT-3 and GPT-4 is the size of the model. GPT-4 has substantially more parameters (1 trillion parameters) than GPT-3 (175 billion parameters)[19]. This increase in parameters enables GPT-4 to learn and retain a wider range of information, and is reported to perform better on a variety of other exams[14, 20].
2. Training data: GPT-4 is trained on a more extensive and diverse dataset than GPT-3. This dataset includes a broader range of sources and languages, which may allow GPT-4 to provide more accurate and relevant information in response to user queries. GPT-3 was trained on 17 gigabytes of data while GPT-4 was trained on 45 gigabytes of data.
3. Generalization and reasoning: GPT-4 exhibits better performance than GPT-3 on benchmarks aimed at measuring generalization and reasoning abilities[21]. For example, GPT-4 outperforms GPT-3 on a variety of logic based multi-choice machine reading comprehension tasks [20, 21]. Thus, we expect GPT-4 to perform better on the ISCD exam.

### Statistical considerations

A practicing clinical physician investigator with a specialty in rheumatology and CCD certification evaluated each question on the practice exam, placed each question into one of the 5 subject categories, and scored the correctness of the responses from of 0 to 4 using the following scale: 0=nonsensical or totally wrong, 1=mostly wrong but in the right field or topic, 2=technically wrong but mostly right, 3=correct answer but wrong explanation, and 4=correct answer and explanation. If the GPT outputs matched one of the exam answers, right or wrong, then the GPT response was noted. If the GPT output was either ambiguous (more than one answer suggested), or nonsensical (response not relevant to answer the question) then a random number generator was used to make a choice. Each response was also evaluated for word count and simple statistics (mean, standard deviation, minimum, maximum) were calculated by GPT version and question category. The correctness and performance of GPT-3 and GPT-4 were compared using a confusion matrix.

## Results

Only questions with a correctness score of 3 or 4 were considered correct. We found that GPT-3 answered 62 of the 100 questions correctly and received an ISCD score 289. GPT-4 answered 82 of the 100 questions correctly and received an ISCD score of 342. There were 58 questions both algorithms got right and 15 both got wrong. See (Table 1).

**Table 1.** Cross Tabulation table between GPT-3 and GPT-4 answers.

|  | *GPT-3*<br>*Correct* | *GPT-3*<br>*Incorrect* |
| --- | --- | --- |
| *GPT-4*<br>*Correct* | 58 | 23 |
| *GPT-4*<br>*Incorrect* | 4 | 15 |

Considering only the correct questions, the average response lengths were close to five times longer for GPT-4 than for GPT-3. See (Table 2). While both had cases where the responses were as short as 1–2-word answers, the maximum word count for a GPT-4 response was 164 versus 56 words for GPT-3.

**Table 2.** Word Count Comparison between GPT-3 and GPT-4 answer responses.

|  | *GPT-3* | *GPT-4* |
| --- | --- | --- |
| *Total Word Count* | 1156 | 5001 |
| *Average Words* | 11.6 | 50.0 |
| *SD* | 19.0 | 43.6 |
| *Min* | 1 | 2 |
| *Max* | 56 | 164 |

When looking at the percentage of answers correct by category, GPT-3 scored the highest on “Overview of Low Bone Mass and Osteoporosis” questions and had the lowest scores on “Imaging Technology in Bone Health”. See (Table 3). Compared to GPT-3, GPT-4 scored higher on all categories with percent right by category ranging from 65% to 91%.

**Table 3.** Breakdown of Answer Accuracy by topic categories for GPT-3 and GPT-4.

|  | **Correctness Score<sup>a</sup>** | **Overview of Low Bone Mass and Osteoporosis** | **Imaging Technology in Bone Health** | **Clinical Application of Bone Densitometry** | **Prevention and Risk Assessment** | **Treatment of Low Bone Mass and Osteoporosis** | **+ Total** |
| --- | --- | --- | --- | --- | --- | --- | --- |
| **GPT-3** | 0 | 5 | 8 | 6 | 3 | 1 | 23 |
|  | 1 | 5 | 4 | 2 | 0 | 1 | 12 |
|  | 2 | 1 | 0 | 0 | 1 | 0 | 2 |
|  | 3 | 1 | 0 | 0 | 0 | 0 | 1 |
|  | 4 | 27 | 11 | 15 | 7 | 2 | 62 |
|  | Correct (%) | 72 | 43 | 65 | 64 | 50 |  |
| **GPT-4** | 0 | 0 | 3 | 0 | 0 | 0 | 4 |
|  | 1 | 2 | 5 | 2 | 1 | 0 | 10 |
|  | 2 | 2 | 0 | 1 | 0 | 1 | 4 |
|  | 3 | 0 | 0 | 0 | 0 | 0 | 0 |
|  | 4 | 35 | 15 | 19 | 10 | 3 | 82 |
|  | Correct (%) | 87 | 65 | 83 | 91 | 75 |  |

<sup>a</sup> 0=nonsensical or totally wrong, 1=mostly wrong but in the right field or topic, 2=technically wrong but mostly right, 3=correct answer but wrong explanation, 4=correct answer and explanation

When looking at correctness, GPT-3 had 85% of its responses either correct or nonsensically wrong 62% and 23%, respectively. In other words, majority of responses were either totally right or totally wrong. See Table 3. This was less true for GPT-4 with only 4% of its responses being totally wrong. There were 5 times that GPT-3 gave an ambiguous answer where a random number generator had to be used, while with GPT-4 this only happened once.

Beyond our numerical analysis, we found that there were nuances in how GPT responded based on the version used that are not easily reflected in raw exam performance. For example, there was an additional degree of fluency and explanation in the responses of GPT-4 when compared to GPT-3. For example, when asked the question, “What area of the lumbar spine is most likely to experience injury?”, GPT-3 responded briefly with “The area of the lumbar spine most likely to experience injury is the L4-L5 region.”, while GPT-4 gave more explanation by stating “The most commonly injured area of the lumbar spine is the L4-L5 level, which is the fourth and fifth lumbar vertebrae. This area is particularly vulnerable to injury due to its location and the amount of stress and load it bears during everyday activities like lifting, bending, and twisting. Injuries to the L4-L5 level can cause lower back pain, sciatica, and other nerve-related symptoms”. Both versions of GPT did well in factual questions such as “What does WHO stand for?” but struggled on questions with some ambiguity such as “What occurs as the rotation of the femoral neck increases or decreases?” The examinee needs to assume an initial position/orientation of the femur to generate a meaningful response. The investigators assume that the question means for the femur to be positioned with the femur neck parallel to the table as is recommended. In this position, the femur neck area is maximized. Rotation in either direction would decrease the bone area increasing the bone mineral density.

Interestingly, one of the questions that GPT-3 got correct but GPT-4 struggled with was a simple logical problem with “How many ulnas are in the human body”. GPT-3 answered correctly with “2”, but GPT-4 responded that there is only one ulna in the human body. Such a question would be very easy for a human to interpret. While we only have one ulna in the forearm, since we have two arms, there are two total ulnas in the human body.

## Discussion

We found that the latest version of the popular GPT program (GPT-4) outperformed an earlier release (GPT-3) in all clinical testing categories in the medical subspecialty of reading and interpreting bone densitometry exams. We also found that the latest version, GPT-4, included more thorough explanations of responses. If the test had been an actual ISCD CCD exam, and GPT-4 had been a human examinee, it would have passed with 82% of the questions correct and an ISCD score of 342. On the contrary, its predecessor, GPT-3, would not have passed. This study demonstrates that modern LLMs increasingly contain specialized knowledge of bone densitometry, and have already passed the threshold used to certify human physicians.

This is the first study that we know of to evaluate LLMs in bone densitometry. However, we do have comparative exam results from the literature on human examinees. Wu et al. [22] analyzed 732 examinees in Taiwan (621 male) over a 7-year period. The average pass rate was 75% with the only predictor of pass rate being age and clinical experience of the examinee in multivariate analysis. Older physicians and/or those with more experience in the field were more likely to pass than younger less experienced examinees.

GPT-4 gave on average 50-word explanations for its responses that allowed highly trained observers to understand the rationale and explanation for the right and wrong responses, close to 5 times the length of the responses from GPT-3. This step is critical to allow a clinician to make an informed decision, and to mitigate automation bias in clinical decision-making. While ChatGPT is a highly advanced language model with a wide range of knowledge, including medical topics, it is important to approach using it as a medical resource with caution. GPT-3 had a training data cutoff date of September 2021. As a result, it is not up-to-date on medical advancements, research, or guidelines changes past that date. For example, the ISCD had a positions development conference in March 2023 that added to and changed their extensive positions statements. Neither version of GPT would have yet learned about these changes.

Other popular LLMs are likely to perform similarly. Meta’s LaMDA is based on the GPT 3.5 architecture and trained on additional data [23]. Google’s Bard is based on LaMDA and trained on more data [24]. The training data and details of these models are not public, so we are unable to predict their performance on the ISCD exam, but our methods could be used to measure the performance empirically LLMs are prone to hallucinations. LLMs can and do generate responses even when it has had little exposure to the topic in the training data [25], and generate a low confidence response. In this case, we may refer to this as the human-equivalent to a “best guess”. Understanding the confidence in each response is critical for using GPT as a medical aid and is an important area for future research.

Passing the CCD examination is a single measure of knowledge of bone densitometry and its applications in osteoporosis care. There are many different aspects that need to be taken into account in the complex clinical care of an osteoporosis patient in real world settings, including individualized patient history and comorbidities, and insurance coverage [26]. Therefore, GPT, and other LLMs at the current state of development are not able to wholly substitute physician care and clinical judgment. Specific applications of LLMs may likely require further training based on expert input to achieve reproducible, high-quality and high-confidence clinical outcomes in the future [27]. It may be ideal to develop deep learning models to complement the clinician’s training and judgment in clinical care, and to simplify and increase efficiency in clinical workflows [28]. Nevertheless, significant guardrails need to be in place to ensure safe patient care, as well as dissemination of accurate information to patients. With further improvements and data, LLMs may provide supplementary input to augment basic understanding of bone densitometry topics and other systematical clinical tasks.

## Limitations

A limitation of our study is that we used a practice test instead of the actual certification exam. While much of the content should be similar between exams, there is still a possibility that the exam used in this study could have questions that are not representative of the official examination. The authors feel that the current study exemplifies the possibilities and potential of GPT in this subspecialty, and may provide motivation for others to explore GPT augmented clinical care.

## Conclusions

If this had been an actual certification exam, GPT-4 would now have a CCD suffix to its name even after being trained using general internet knowledge. Clearly, more goes into physician training than can be captured in this exam. However, GPT algorithms may prove to be valuable physician aids in the diagnoses and monitoring of osteoporosis and other diseases. We conclude that the current version of GPT-4 has sufficient specialized knowledge of bone densitometry to pass the ISCD CCD exam. We anticipate that successive versions of GPT algorithms will be useful to augment clinical decision making and expedite clinical care.

## Data Availability

All data produced in the present study are available upon reasonable request to the authors.

## Abbreviations

AI: artificial intelligence
CCD: certified clinical densitometrist
GPT: generative pre-trained transformer
ISCD: international society of clinical densitometry
LLM: large language model
NLP: natural language processing

